# Reassessing Asymmetry Reduction in Psychosis: Cingulate Folding and Gyrification Covariance in Patients with Auditory Hallucinations

**DOI:** 10.1101/2024.11.07.24316889

**Authors:** Shun-Chin Jim Wu, Héloïse de Vareilles, Samantha C. Mitchell, Atheer Al-Manea, Jane Garrison, Michail Mamalakis, Jon S. Simons, Arnaud Cachia, Jean-François Mangin, Stener Nerland, Lynn Mørch-Johnsen, Ingrid Agartz, John Suckling, Graham K. Murray

**Affiliations:** Department of Psychiatry, University of Cambridge, UK; Department of Psychology, University of Cambridge, UK; Centre for Human Brain Health, School of Psychology, University of Birmingham, Birmingham, UK; Université Paris Cité, LaPsyDÉ, CNRS, Paris, France; Université Paris Cité, Institut de Psychiatrie et Neurosciences de Paris (IPNP), INSERM, F-75014 Paris, France; Paris-Saclay University, CNRS, CEA, BAOBAB, NeuroSpin, Gif-sur-Yvette, France; Division of Mental Health and Substance Abuse, Diakonhjemmet Hospital, Oslo, Norway; Division of Mental Health and Addiction, Institute of Clinical Medicine, University of Oslo, Oslo, Norway; Department of Psychiatry, Østfold Hospital, Grålum, Norway; Department of Clinical Research, Østfold Hospital, Grålum, Norway; Department of Psychiatric Research, Diakonhjemmet Hospital, Oslo, Norway; Centre for Psychiatry Research, Department of Clinical Neuroscience, Karolinska Institute and Stockholm Health Care Services, Stockholm Region, Stockholm, Sweden; Cambridgeshire and Peterborough NHS Trust, Cambridgeshire, UK

**Keywords:** Paracingulate sulcus, Asymmetry, Auditory hallucinations, Psychosis continuum, Structural covariance

## Abstract

**Background and Hypothesis:** Prior research links a shorter paracingulate sulcus (PCS) to hallucinations in schizophrenia, but its symmetry hemispheric specificity and relevance to bipolar disorders remain unclear. We hypothesized that reduced PCS asymmetry and interhemispheric gyrification covariance in salience and auditory networks are associated with lifetime auditory hallucinations (AH) in psychotic spectrum disorders.

**Study Design:** We compared patients with and without AH, and healthy controls, focusing on PCS asymmetry in five ordinal classes, sulcal length and depth, and interhemispheric gyrification covariance.

**Study Results:** Among 351 patients with schizophrenia or bipolar spectrum disorders (SSD/BSD), 194 (55.3%) had AH, compared to 157 without and 278 healthy controls. We found no significant PCS class asymmetry between hemispheres (V = 6648.5, *P* = .097) and decreased leftward asymmetry in PCS length (F(2,626) = 3.19, *P* = .013) in patients with AH, compared with those without and healthy controls. Compared to patients without AH, those with AH showed increased gyrification covariance in the auditory network (F(2,625) = 42.5, *P* < .001). In the salience network, patients with SSD and AH had increased covariance (F(2,625) = 299, *P* < .001), while patients with BSD and AH displayed decreased covariance (F(2,625) = 102, *P* < .001).

**Conclusions:** This study, featuring the largest cohort to date, links the AH trait to replicable reduced leftward PCS asymmetry and altered interhemispheric covariance in psychotic spectrum disorders, supporting theories of reduced asymmetry and altered brain network coordination as part of the mechanistic pathway to psychosis.

## Introduction

Reduced brain asymmetry has been proposed as one of etiological hypotheses for psychosis^1,2^, supported by deviations in the anterior cingulate cortex and insula observed in both patients with schizophrenia and bipolar disorder compared to healthy individuals^3–5^. Although earlier studies were constrained by limited sample sizes, recent large-scale research has identified alternated brain asymmetry in schizophrenia, particularly in the cortical thickness of the rostral anterior cingulate and middle temporal gyrus, compared to heathy individuals^6^.

Cortical thickness and volume fluctuate significantly during maturation, but the topological measure like cortical folding around the anterior cingulate cortex, bordered by the paracingulate sulcus (PCS), is established prenatally and remains stable throughout life^7^. In addition, the cortical topology of the medial frontal cortex including the PCS has been associated with a liability to hallucinations in schizophrenia spectrum disorders^8–11^, and executive function and reality monitoring, psychological processes that may, when impaired, contribute to psychotic symptom pathogenesis^12–14^. The anterior cingulate cortex is consistently related to PCS, typically exhibiting rightward asymmetry in the ACC and leftward asymmetry in the PCS^15–18^. Therefore, the PCS may serve as a stable marker for susceptibility to hallucinations throughout life^19–21^.

However, since the presence and morphology of the PCS vary widely across individuals and hemispheres^7^, findings on laterality deviations are mixed. A shorter left PCS is often observed in psychotic disorders, particularly in those with hallucinations, but some studies report right sided or bilateral deviations^8–11,22–28^. Asymmetry measures may help reconcile these inconsistencies, as the extent of deviations in asymmetry have been linked to the severity or liability of psychotic symptoms^5,11,29^. Therefore, asymmetry of the medial frontal lobe has been suggested as a transdiagnostic biomarker for the psychosis continuum, indicating a shared risk feature across the schizophrenia and bipolar disorder spectrum^1,3–5^.

Topological measures can also be assessed through structural covariance, which reflects the coordination between brain regions or networks^30–33^. Many previous studies have relied on expert anatomical annotation of folding patterns; however, brain surface topology can also be quantified using measures such as local gyrification^34^. We have previously reported alterations in the covariance of gyrification indices both within and between regions in the salience and auditory networks, suggesting that perinatal alterations in developmental trajectories may be linked to the experience of hallucinations in early adulthood^11^. Given that asymmetry deviations influence cortical topology in both hemispheres, these effects may be evident in the interhemispheric structural covariance of gyrification in the PCS and related networks.

Inconsistent findings in this field may stem from several limitations in previous studies: modest sample sizes, focus only on schizophrenia spectrum disorders, single measurement method (either categorical or continuous), and a reliance on bilateral structural covariances, which are obscured by unilateral within-network covariance due to their close geodesic distance^11,16,35^. To overcome these limitations and explore the potential of PCS morphological asymmetry as a transdiagnostic biomarker for auditory hallucinations in schizophrenia and bipolar spectrum disorders, this study aims to examine PCS asymmetry using both categorical and continuous methods. Additionally, it incorporates interhemispheric structural covariance analysis of gyrification in the PCS and extended salience and auditory networks, using a large cohort (N = 629) comprising patients with schizophrenia and bipolar spectrum disorders, along with healthy controls.

Theoretically, we seek to provide improved evidence of topological coordination within the PCS and the salience and auditory networks in the early co-development of structures that implicate the risk for schizophrenia or bipolar disorder, particularly regarding the hallucinations experience as a symptom of psychosis. These brain regions and networks are central to the theory of misattributed reality monitoring and/or inner speech as mechanisms for auditory hallucinations^36^. Our specific hypotheses are that patients with a history of lifetime auditory hallucinations (lifetime AH) exhibit decreased leftward asymmetry and interhemispheric covariance in the PCS, but increased covariance within and between the salience and auditory networks.

## Methods

This study compared PCS asymmetry using a categorical rating scale and continuous metrics of sulcal morphology, alongside interhemispheric structural covariance of gyrification, between patients with and without lifetime AH among psychotic spectrum disorders, and healthy controls. Additionally, interhemispheric structural covariance within and between salience and auditory networks across these groups were investigated. The overall study design is outlined in Figure 1. To further discern potential differences between subgroups (schizophrenia spectrum disorders and bipolar spectrum disorders), all analyses were also conducted within each subgroup (stratified analysis).

**Figure 1.**
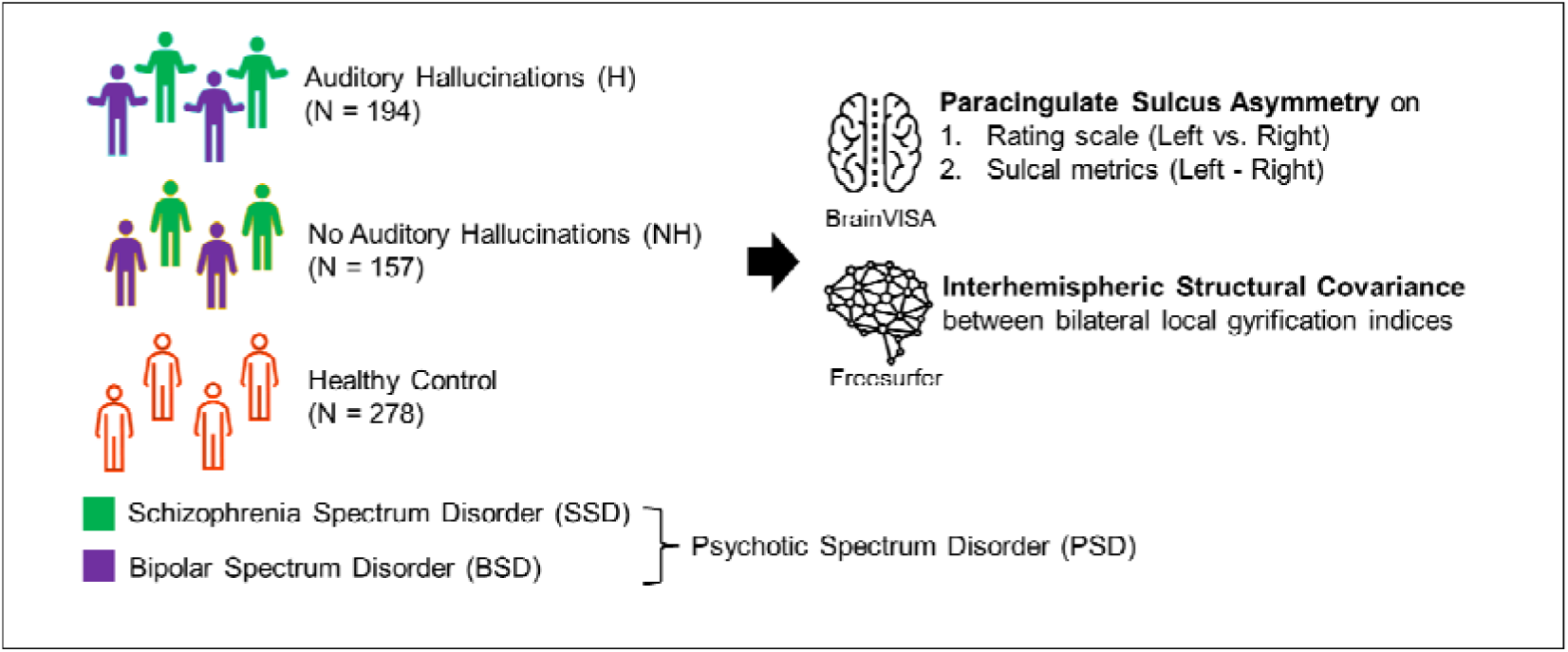
Overview of the Study. This study examined patients with psychotic spectrum disorders (PSD), including schizophrenia spectrum disorders (SSD) and bipolar spectrum disorders (BSD), who experience lifetime auditory hallucinations (N = 194), compared to those without auditory hallucinations (N = 157), as well as healthy controls (N = 278) from the Thematically Organized Psychosis study. We investigated paracingulate sulcus asymmetry using a categorical rating scale and continuous metrics, along with interhemispheric structural covariance in gyrification within and between salience and auditory networks.

### Dataset

The study utilized structural T1-weighted MRI images obtained from the Thematically Organized Psychosis study conducted at the University of Oslo and the Oslo University Hospital, focusing on patients diagnosed with psychotic disorders^37,38^. Images were acquired using a 1.5 Tesla Siemens MAGNETOM Sonata scanner (Siemens Medical Solutions, Erlangen, Germany) equipped with a standard head coil. After obtaining a conventional 3-plane localizer, two sagittal T1-weighted volumes were acquired using the magnetization prepared rapid gradient echo (MPRAGE) technique with the Siemens tfl3d1_ns pulse sequence. The acquisition parameters were as follows: echo time (TE) = 3.93 ms, repetition time (TR) = 2730 ms, inversion time (TI) = 1000 ms, flip angle = 7°, field of view (FOV) = 24 cm, voxel size = 1.33×0.94×1 mm³, and 160 partitions. To enhance the signal-to-noise ratio, the two acquired MPRAGE volumes were first registered with the rigid-body registration and then averaged. Notably, there were no significant changes to the scanner or related instruments during the study’s image acquisition period.

In total, 629 participants were included, comprising 351 patients diagnosed with schizophrenia or bipolar spectrum disorder based on the Structured Clinical Interview for DSM-IV (SCID-IV), and 278 healthy controls. The presence or absence of lifetime auditory hallucinations in patients was determined using the item B16 from the SCID-IV. Patients were combined into a category termed psychotic spectrum disorders, with 194 patients with lifetime AH (55.3%) and 157 patients without (44.7%). Specifically, there were 194 patients with schizophrenia spectrum disorders, categorized as follows: schizophrenia (N = 145), schizoaffective disorder (N = 28), and schizophreniform disorder (N = 21). Additionally, there were 157 patients with bipolar spectrum disorders, divided into bipolar I disorder (N = 98), bipolar II disorder (N = 50), and bipolar disorder not otherwise specified (N = 9). Around 74.7% of patients with schizophrenia spectrum disorders and 31.1% of patients with bipolar spectrum disorders reported lifetime AH. Demographic and clinical profiles are provided in Table 1.

**Table 1.**
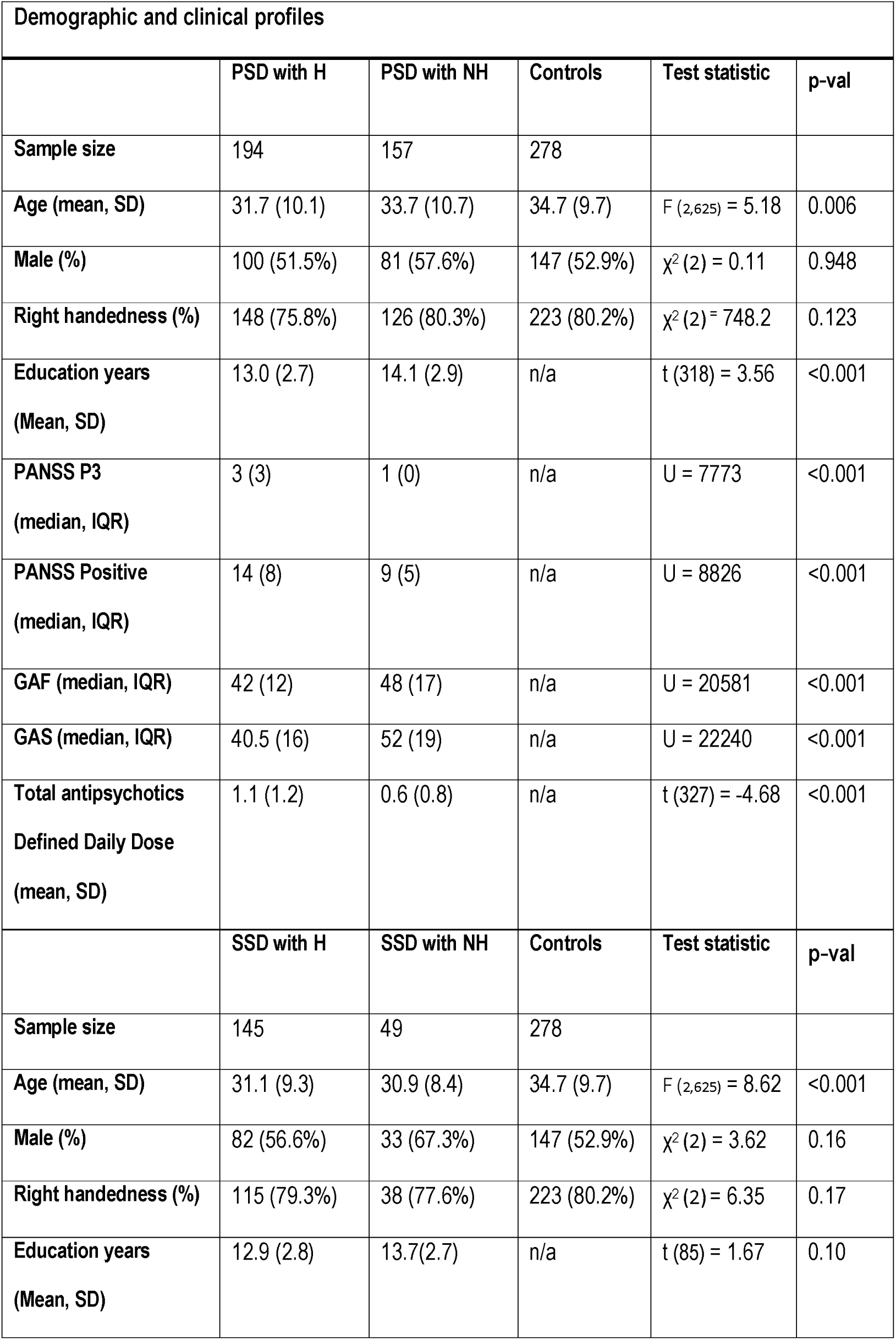
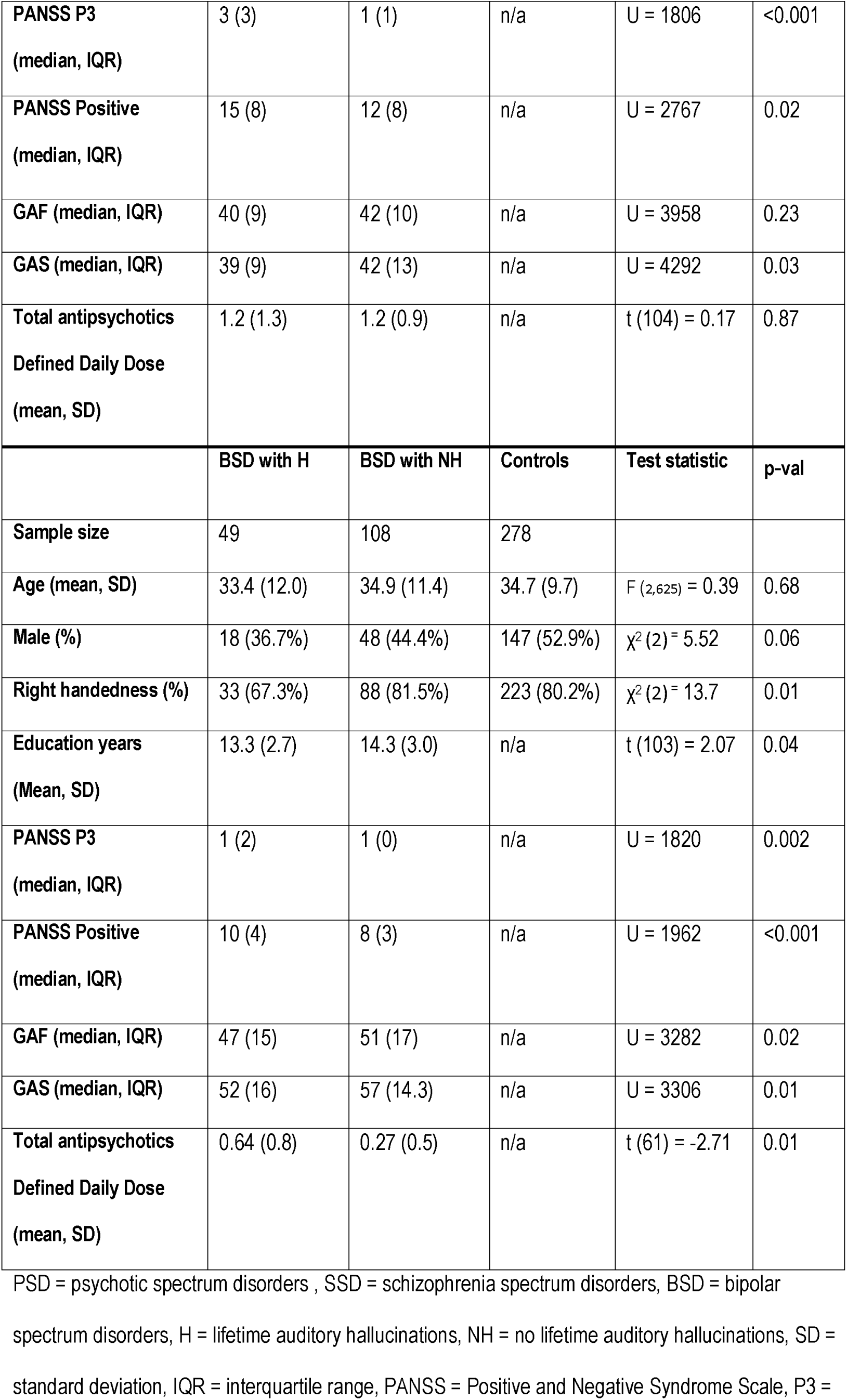
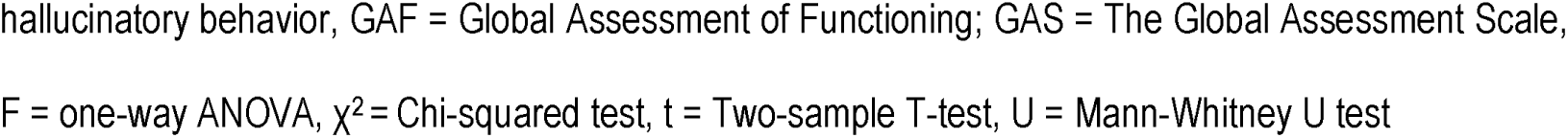
Demographics and Clinical Information.

| Demographic and clinical profiles |  |  |  |  |  |
| --- | --- | --- | --- | --- | --- |
|  | PSD with H | PSD with NH | Controls | Test statistic | p-val |
| Sample size | 194 | 157 | 278 |  |  |
| Age (mean, SD) | 31.7 (10.1) | 33.7 (10.7) | 34.7 (9.7) | $F_{(2,625)} = 5.18$ | 0.006 |
| Male (%) | 100 (51.5%) | 81 (57.6%) | 147 (52.9%) | $\chi^2 (2) = 0.11$ | 0.948 |
| Right handedness (%) | 148 (75.8%) | 126 (80.3%) | 223 (80.2%) | $\chi^2 (2) = 748.2$ | 0.123 |
| Education years<br>(Mean, SD) | 13.0 (2.7) | 14.1 (2.9) | n/a | $t (318) = 3.56$ | <0.001 |
| PANSS P3<br>(median, IQR) | 3 (3) | 1 (0) | n/a | $U = 7773$ | <0.001 |
| PANSS Positive<br>(median, IQR) | 14 (8) | 9 (5) | n/a | $U = 8826$ | <0.001 |
| GAF (median, IQR) | 42 (12) | 48 (17) | n/a | $U = 20581$ | <0.001 |
| GAS (median, IQR) | 40.5 (16) | 52 (19) | n/a | $U = 22240$ | <0.001 |
| Total antipsychotics<br>Defined Daily Dose<br>(mean, SD) | 1.1 (1.2) | 0.6 (0.8) | n/a | $t (327) = -4.68$ | <0.001 |
|  | SSD with H | SSD with NH | Controls | Test statistic | p-val |
| Sample size | 145 | 49 | 278 |  |  |
| Age (mean, SD) | 31.1 (9.3) | 30.9 (8.4) | 34.7 (9.7) | $F_{(2,625)} = 8.62$ | <0.001 |
| Male (%) | 82 (56.6%) | 33 (67.3%) | 147 (52.9%) | $\chi^2 (2) = 3.62$ | 0.16 |
| Right handedness (%) | 115 (79.3%) | 38 (77.6%) | 223 (80.2%) | $\chi^2 (2) = 6.35$ | 0.17 |
| Education years<br>(Mean, SD) | 12.9 (2.8) | 13.7(2.7) | n/a | $t (85) = 1.67$ | 0.10 |
| **PANSS P3**<br>(median, IQR) | 3 (3) | 1 (1) | n/a | U = 1806 | <0.001 |
| **PANSS Positive**<br>(median, IQR) | 15 (8) | 12 (8) | n/a | U = 2767 | 0.02 |
| **GAF (median, IQR)** | 40 (9) | 42 (10) | n/a | U = 3958 | 0.23 |
| **GAS (median, IQR)** | 39 (9) | 42 (13) | n/a | U = 4292 | 0.03 |
| **Total antipsychotics**<br>**Defined Daily Dose**<br>(mean, SD) | 1.2 (1.3) | 1.2 (0.9) | n/a | t (104) = 0.17 | 0.87 |
|  | **BSD with H** | **BSD with NH** | **Controls** | **Test statistic** | **p-val** |
| **Sample size** | 49 | 108 | 278 |  |  |
| **Age (mean, SD)** | 33.4 (12.0) | 34.9 (11.4) | 34.7 (9.7) | F (2,625) = 0.39 | 0.68 |
| **Male (%)** | 18 (36.7%) | 48 (44.4%) | 147 (52.9%) | $\chi^2$ (2) = 5.52 | 0.06 |
| **Right handedness (%)** | 33 (67.3%) | 88 (81.5%) | 223 (80.2%) | $\chi^2$ (2) = 13.7 | 0.01 |
| **Education years**<br>(Mean, SD) | 13.3 (2.7) | 14.3 (3.0) | n/a | t (103) = 2.07 | 0.04 |
| **PANSS P3**<br>(median, IQR) | 1 (2) | 1 (0) | n/a | U = 1820 | 0.002 |
| **PANSS Positive**<br>(median, IQR) | 10 (4) | 8 (3) | n/a | U = 1962 | <0.001 |
| **GAF (median, IQR)** | 47 (15) | 51 (17) | n/a | U = 3282 | 0.02 |
| **GAS (median, IQR)** | 52 (16) | 57 (14.3) | n/a | U = 3306 | 0.01 |
| **Total antipsychotics**<br>**Defined Daily Dose**<br>(mean, SD) | 0.64 (0.8) | 0.27 (0.5) | n/a | t (61) = -2.71 | 0.01 |

PSD = psychotic spectrum disorders, SSD = schizophrenia spectrum disorders, BSD = bipolar
spectrum disorders, H = lifetime auditory hallucinations, NH = no lifetime auditory hallucinations, SD =
standard deviation, IQR = interquartile range, PANSS = Positive and Negative Syndrome Scale, P3 =
hallucinatory behavior, GAF = Global Assessment of Functioning; GAS = The Global Assessment Scale, F = one-way ANOVA, $\chi^2$ = Chi-squared test, t = Two-sample T-test, U = Mann-Whitney U test

### Image preprocessing

To accurately delineate individual sulcus for visual classification and manual labelling, we employed 3-D visualization using BrainVISA. For precise calculation of regional gyrification, vertex-wise surface analysis was conducted with Freesurfer. Consistent with our previous research on hallucinatory behaviors in schizophrenia^11^, we chose to use the same two software tools to ensure methodological consistency.

- **BrainVISA:** Due to the variability of the PCS, automatic labelling of the PCS is not implemented in BrainVISA’s Morphologist automatic sulcal recognition pipeline (version 5.0.4)^39,40^. Instead, visual rating and manual labelling of the PCS were performed for each participant’s hemispheres. Prior to rating and labelling, T1-weighted MR images underwent preprocessing using the Morphologist pipeline^41^, involving grey and white matter segmentation, mesh creation, and cortical folds graph generation. PCS categorization and labelling were independently conducted on the unlabeled cortical folds graph, without knowledge of the diagnosis or sulcal patterns in the opposite hemisphere. The PCS was visually classified based on its presence and morphology, particularly focusing on its length and width, in accordance with the Paracingulate Sulcus Classification Protocol developed by our colleagues^42^. The PCS was also manually labelled according to the Paracingulate Sulcus Labelling Protocol also developed by our colleagues^43^. Following this manual labelling, metrics including sulcal length and depth were automatically generated and extracted. Emphasizing sulcal length and depth aligns with previous literature presented in the introduction. Estimated intracranial volume and hemispheric brain volume were also derived by BrainVISA to be used as covariates in subsequent analysis.
- **Freesurfer:** After preprocessing T1-weighted MR images with “recon-all-all” in Freesurfer (version 6.0.0), the local gyrification index (LGI), a quantitative measure representing the amount of embedded cortex within each cortical vertex, was calculated for gyrification. One healthy control subject was excluded from subsequent structural covariance analyses due to processing failure (resulting in N = 628). Interhemispheric structural covariance for the PCS, salience and auditory networks was established by resampling LGI maps from vertices to regions on the Human Connectome Project-MMP1.0 multimodal surface-based anatomical atlas^44^. Specifically, we focused on the PCS, incorporating indices within 24dd, 24dv, SCEF, p32pr, a32pr, d32, p32, 8BM, and 9m. Similarly, for the salience network, the regions FEF, 5mv, 23c, SCEF, 6ma, p24pr, 33pr, a24pr, p32pr, 6r, 46, 9-46d, 43, PFcm, PoI2, FOP4, MI, AVI, AAIC, FOP1, FOP3, FOP2, PFop, PF, PoI1, FOP5, and a32pr were included. The auditory network comprised the regions A1, PSL, STV, OP4, OP1, OP2-3, 52, RI, TA2, STGa, PBelt, A5, STSda, STSdp, STSvp, TPOJ1, Ig, MBelt, LBelt, A4, and STSva. Lastly, estimated intracranial volume was also calculated by Freesurfer to be used as a covariate in subsequent analyses of structural covariances in gyrification.

### Paracingulate sulcus asymmetry

1. **Categorical rating scale:** To classify the PCS more precisely than the classical three-class system^45^, the PCS on the unlabeled cortical folds graph was visually rated into five ordinal classes based on the following criteria (see Figure 2):

Class 1: a complete absence of folding, with minimal or no sulcal elements observed dorsal to the cingulate sulcus, or the presence of folding dorsal to the cingulate sulcus that do not parallel the cingulate sulcus.
Class 2: the presence of sparse sulcal paralleling to the cingulate sulcus but failing to globally outline a complete PCS. The gaps between sulcal elements are more noticeable than the sulcal elements themselves.
Class 3: an interrupted PCS, where sulcal elements outline a complete PCS but with missing portions or limited spatial extension. The elements of the potential full PCS are more noticeable than the gaps between the elements.
Class 4: a full-length PCS extending from the extension of the genu of the corpus callosum to the vertical anterior commissure line, shallower than the cingulate.
Class 5: an almost cingulate-like PCS, which is deep and continuous and could be mistaken for the cingulate sulcus To examine the asymmetry of the PCS classes, the presence of asymmetry between bilateral PCS classes (left PCS class vs. right PCS class) within each group was initially assessed. The Wilcoxon signed-rank test (two-sided) was applied to compare left and right PCS classes pairwise within each group, using the stats package (version 3.6.2) in R (version 4.2.3). Asymmetry was considered absent if the null hypothesis of no significant difference between left and right PCS classes was not rejected, indicated by a p-value of 0.05 or higher.

**Figure 2.**
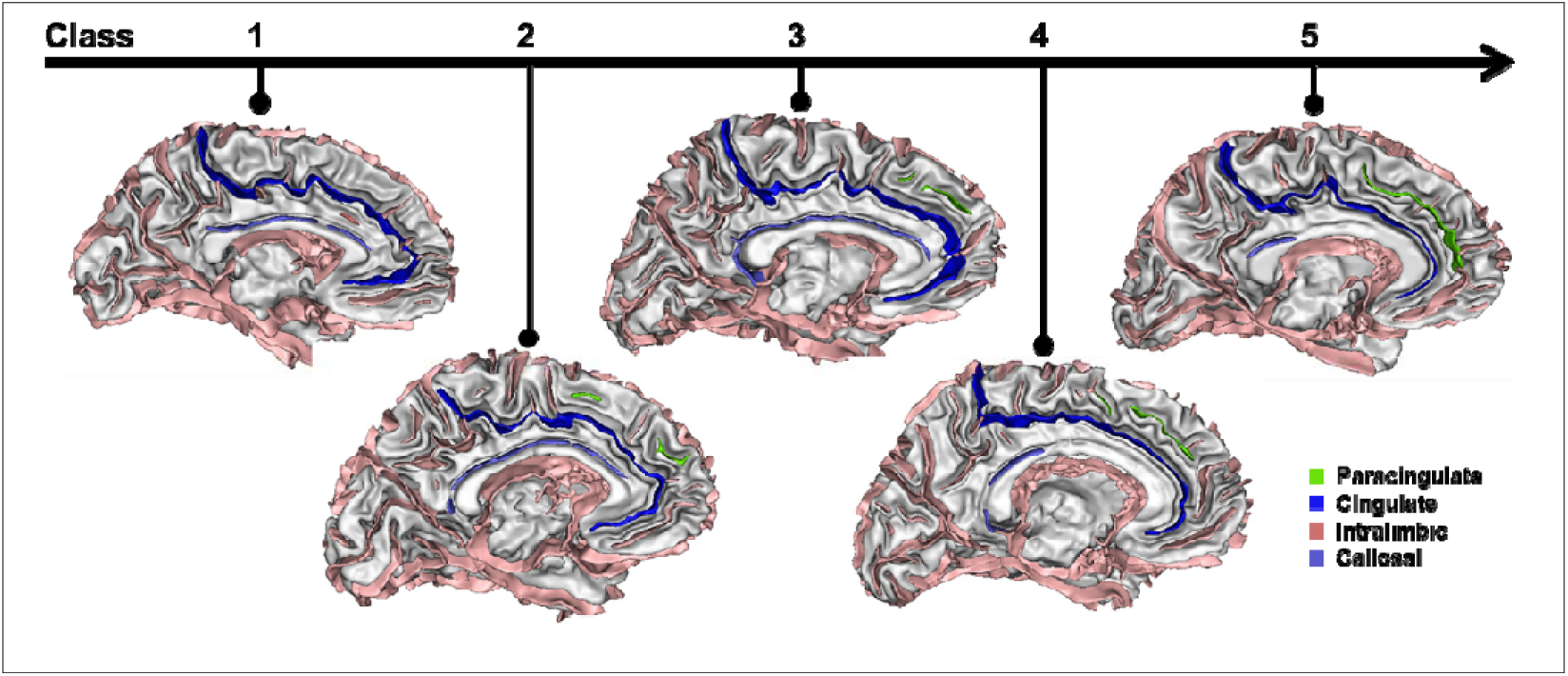
Paracingulate Sulcus Categorical Rating Scale. The paracingulate sulcus, highlighted in light green, can be visually classified into five ordinal classes using the BrainVisa software. The rating scale ranges from no visible sulcus or only elements not parallel to the cingulate sulcus (class 1) to a sparse sulcus with prominent gaps (class 2), an interrupted sulcus (class 3), a full-length sulcus (class 4), and finally, a sulcus paralleling and resembling the cingulate sulcus (in dark blue) with similar depth (class 5). If asymmetry was absent in any group, the Kruskal-Wallis rank sum test was used to compare the differences between bilateral PCS classes (left minus right PCS class) across groups. This test was conducted with the rstatix package (version 0.7.2), assuming that the differences between paired ordinal variables by subtraction are ordinal.
2. **Continuous metrics:** With automatically generated metrics including length and average depth of the PCS in native space, the asymmetry of metrics in the PCS was evaluated by the asymmetry difference (Left − Right) instead of the asymmetry index ((Left − Right) / (Left + Right)), as adopted by our previous work ^11^. This decision aimed to avoid inaccuracies in asymmetry measurement, particularly for cases of unilateral appearance of the PCS, which are not uncommon. The classic asymmetry index may obscure the significance of differences, especially when one side of the metrics is zero, potentially masking variations between large and minimal asymmetry values (e.g., 10 vs. 0.1 mm). For group comparisons, one-way analysis of covariance (ANCOVA) with 5000 permutations was conducted using the permuco package (version 1.1.1) in R. This analysis was controlled for multiple covariates, including age, sex, estimated intracranial volume from BrainVISA, brain volume difference (Left − Right brain volume), the sum of metrics (Left + Right metrics), and broad diagnosis group (schizophrenia spectrum disorders or bipolar spectrum disorders). These covariates were included to mitigate regional asymmetry in the PCS driven by aging, sex difference, head size, global hemispheric asymmetry, total metrics scale, or subgroup diagnosis, respectively^46,47^. If significant differences were found in one-way ANCOVA, post-hoc Tukey Honest Significant Difference (HSD) tests for pairwise comparisons were performed using lmPerm (version 2.1.0) and multcomp packages (version 1.4-25) in R. Additionally, since sexual differences are crucial to Dr. Crow’s reduced asymmetry hypothesis^3^, the effect of the interaction between the lifetime AH and sex on the asymmetry of PCS metrics was also assessed.

### Interhemispheric structural covariance of local gyrification indices

1. **Paracingulate sulcus:** To assess the interhemispheric coordination of the PCS, we computed the interhemispheric structural covariance of LGIs between bilateral PCS regions. This analysis was conducted using partial correlation with Pearson correlations, adjusting for age, sex, estimated intracranial volumes from Freesurfer, and broad diagnosis group (schizophrenia spectrum disorders or bipolar spectrum disorders) with the ppcor package (version 1.1) in R. Subsequently, we retained only significant correlation coefficients that survived after multiple testing corrections with false discovery rate (FDR) for subsequent group comparisons. Group comparisons were then performed using the one-way analysis of variance (ANOVA) with 5000 permutations via the permuco package in R. If a significant difference was observed in one-way ANOVA, post-hoc Tukey HSD tests for pairwise comparisons (two-sided) were conducted using the lmPerm and multcomp packages in R. Lastly, the estimated intracranial volumes calculated by BrainVisa and Freesurfer showed high consistency (Spearman correlation coefficient = 0.951). However, we opted to use the respective values from each tool for greater precision.
2. **Salience and auditory networks:** To explore the interhemispheric coordination within and between salience and auditory networks, we repeated the interhemispheric structural covariance analysis between bilateral salience, auditory, and a combined network encompassing both salience and auditory networks (i.e., the left salience and auditory networks correlated with the right salience and auditory networks). This analysis was carried out using the same aforementioned methods to compare groups.

### Sensitivity analysis

To ensure the comparability with previous studies and the robustness the findings, the sensitivity analysis was conducted as follows:

1. **Using asymmetry index of metrics for PCS:** To align with our prior research^11^, the asymmetry index for PCS metrics was still estimated.
2. **Adding antipsychotics defined daily dose**^48^ **or the Positive and Negative Syndrome Scale (PANSS) positive scores as an additional covariate:** To ensure stability and specificity of findings for auditory hallucination trait, independent of general psychosis status ^49^, the PCS metrics asymmetry analyses were controlled for antipsychotics dose or PANSS positive scores.
3. **Examining unilateral PCS rating scale and metrics:** To facilitate direct comparisons with existing studies used either unilateral categorical rating scales or continuous metrics of the PCS^8–11,22–28,50^ and validate the consistency of the findings, group comparisons with these unilateral rating scales and metrics were also performed.

## Results

### Reduced leftward PCS asymmetry in patients with auditory hallucinations

#### Categorical analysis (PCS class) of asymmetry

When analyzing the PCS classes, a loss of PCS class asymmetry was observed only in patients with psychotic spectrum disorders and lifetime AH, indicated by the absence of a significant difference between bilateral PCS classes (V = 6648.5, Z = 0.12, *P* = .097). In contrast, PCS class asymmetry remained intact in patients without lifetime AH and in healthy controls (V = 13471 and 4610.5, Z = 0.23 and 0.19, *P* = .004 and .005). Subgroup analysis suggested that patients within the bipolar spectrum disorders showed a pronounced loss of PCS class asymmetry (V = 332, Z= 0.12, *P* = .414). However, between-group comparisons revealed no statistically significant difference in PCS class asymmetry (see Table 2).

**Table 2.**
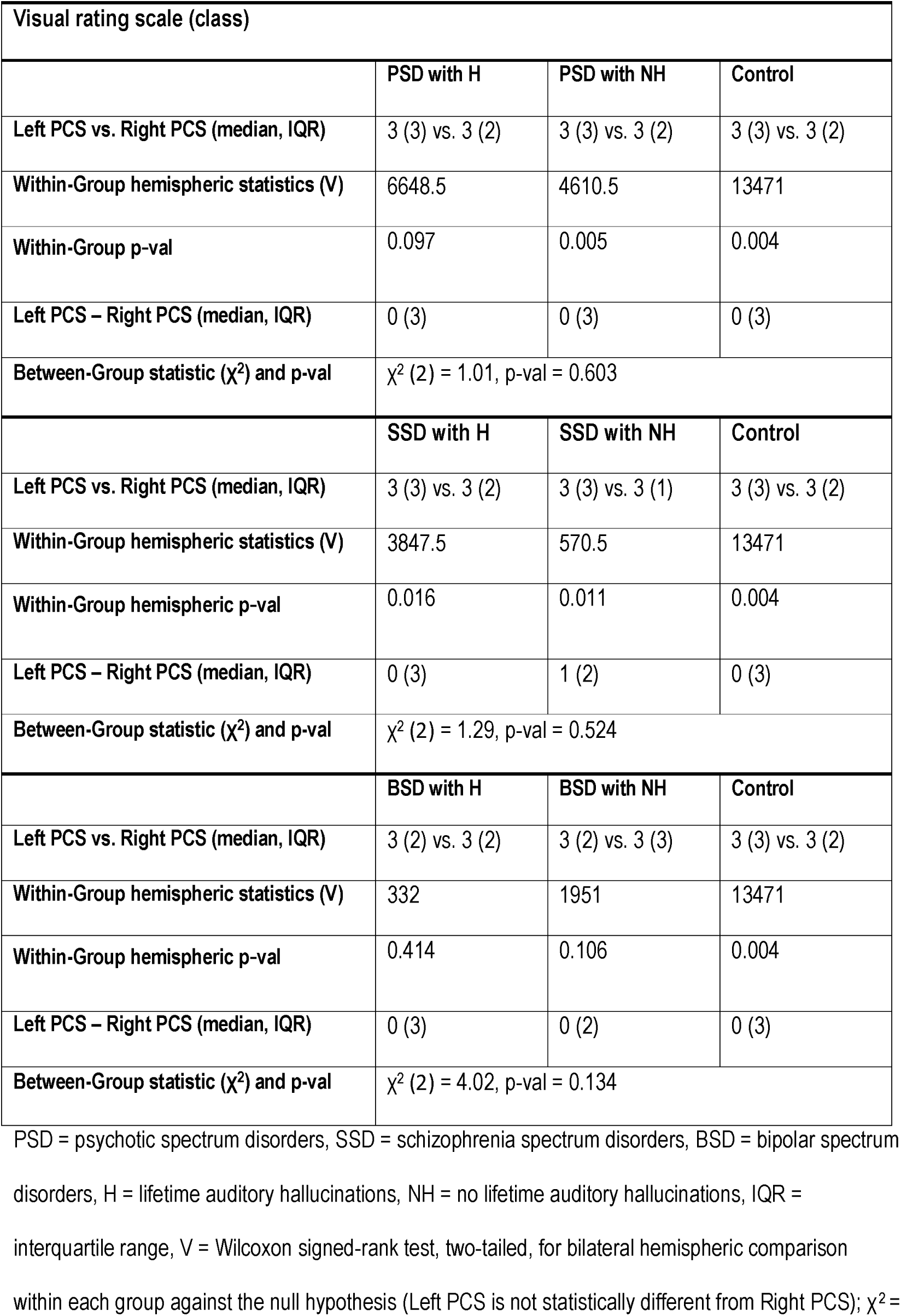
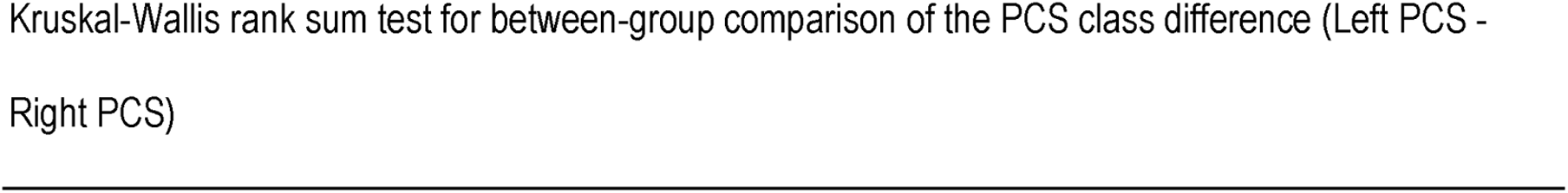
Paracingulate Sulcus (PCS) Rating Scale: Within-Group Hemispheric Comparison and Between-Group Class Difference Comparison.

| Visual rating scale (class) |  |  |  |
| --- | --- | --- | --- |
|  | PSD with H | PSD with NH | Control |
| Left PCS vs. Right PCS (median, IQR) | 3 (3) vs. 3 (2) | 3 (3) vs. 3 (2) | 3 (3) vs. 3 (2) |
| Within-Group hemispheric statistics (V) | 6648.5 | 4610.5 | 13471 |
| Within-Group p-val | 0.097 | 0.005 | 0.004 |
| Left PCS – Right PCS (median, IQR) | 0 (3) | 0 (3) | 0 (3) |
| Between-Group statistic ($\chi^2$) and p-val | $\chi^2$ (2) = 1.01, p-val = 0.603 | | |
|  | SSD with H | SSD with NH | Control |
| Left PCS vs. Right PCS (median, IQR) | 3 (3) vs. 3 (2) | 3 (3) vs. 3 (1) | 3 (3) vs. 3 (2) |
| Within-Group hemispheric statistics (V) | 3847.5 | 570.5 | 13471 |
| Within-Group hemispheric p-val | 0.016 | 0.011 | 0.004 |
| Left PCS – Right PCS (median, IQR) | 0 (3) | 1 (2) | 0 (3) |
| Between-Group statistic ($\chi^2$) and p-val | $\chi^2$ (2) = 1.29, p-val = 0.524 | | |
|  | BSD with H | BSD with NH | Control |
| Left PCS vs. Right PCS (median, IQR) | 3 (2) vs. 3 (2) | 3 (2) vs. 3 (3) | 3 (3) vs. 3 (2) |
| Within-Group hemispheric statistics (V) | 332 | 1951 | 13471 |
| Within-Group hemispheric p-val | 0.414 | 0.106 | 0.004 |
| Left PCS – Right PCS (median, IQR) | 0 (3) | 0 (2) | 0 (3) |
| Between-Group statistic ($\chi^2$) and p-val | $\chi^2$ (2) = 4.02, p-val = 0.134 | | |

PSD = psychotic spectrum disorders, SSD = schizophrenia spectrum disorders, BSD = bipolar spectrum disorders, H = lifetime auditory hallucinations, NH = no lifetime auditory hallucinations, IQR = interquartile range, V = Wilcoxon signed-rank test, two-tailed, for bilateral hemispheric comparison within each group against the null hypothesis (Left PCS is not statistically different from Right PCS); $\chi^2$ =
Kruskal-Wallis rank sum test for between-group comparison of the PCS class difference (Left PCS - Right PCS)

#### Continuous analysis (PCS length) of asymmetry

A significantly decreased leftward asymmetry in PCS length was observed in patients with psychotic spectrum disorders and lifetime AH (F(2,626) = 3.19, *P* = .013 in length). In the post-hoc analysis, compared to patients without a history of auditory hallucinations, those with such a history showed a reduction in PCS length asymmetry by −10.97 mm (Z = −2.53, SE = 4.33, *P* = .028). Subgroup analysis of PCS length asymmetry indicated that patients with bipolar spectrum disorders and lifetime AH exhibited a pronounced shift from leftward to rightward asymmetry, with a transition from a positive to a negative mean asymmetry difference of −4.0 mm. This reduction in PCS length asymmetry in patients with bipolar spectrum disorders and a history of auditory hallucinations was statistically significant compared to healthy controls (−13.67 mm, Z = −2.38, SE = 5.76, *P* = .045) but not when compared to patients without such a history (−11.64 mm, Z = −1.83, SE = 6.35, *P* = .154). Among patients with schizophrenia spectrum disorders, no statistically significant difference in PCS length asymmetry was found across the three groups. There was no statistically significant difference in PCS depth asymmetry between the three groups. Summarized results from the PCS metrics asymmetry are depicted in Figure 3A and Table S1. No significant interaction effect was observed between the lifetime AH and sex on the PCS length or depth asymmetry.

**Figure 3.**
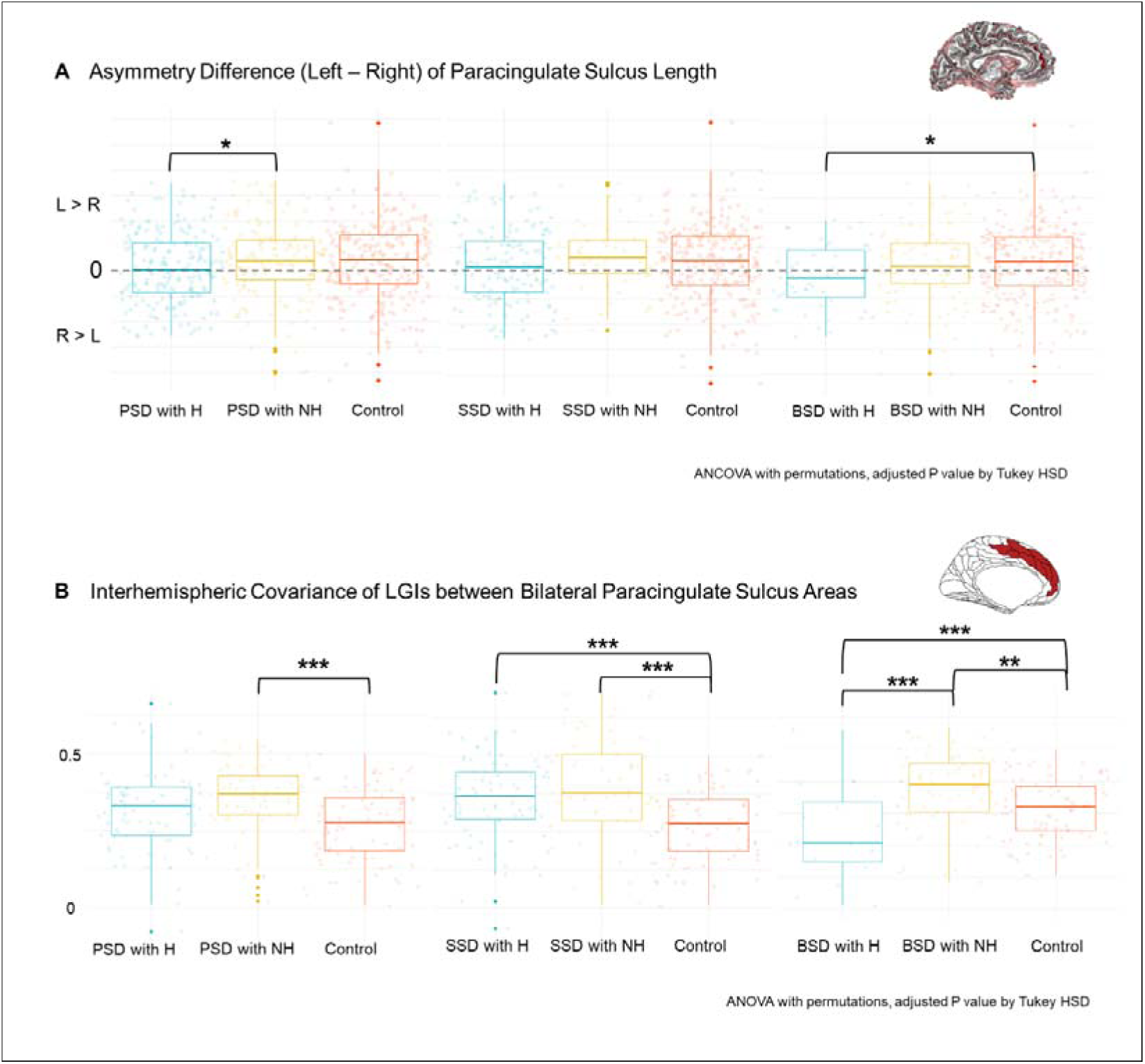
Decreased Leftward Asymmetry and Interhemispheric Covariance in the Paracingulate Sulcus in Patients with Lifetime Auditory Hallucinations. (A) Patients with psychotic spectrum disorders and lifetime auditory hallucinations (PSD with H) exhibited significantly reduced leftward asymmetry in the length of the paracingulate sulcus compared to those without auditory hallucinations (PSD with NH), as indicated by the asymmetry difference (right-left or R − L), assessed using one-way analysis of covariance (ANCOVA) with 5000 permutations (F(2,626) = 3.19, P = .013). Posthoc testing revealed a mean difference in asymmetry of −10.97 mm (SE = 4.33, P = .028). In subgroup analyses, patients with bipolar spectrum disorders and auditory hallucinations (BSD with H) displayed an even more pronounced reduction in leftward asymmetry, indicated by a shift toward rightward asymmetry. (B) Interhemispheric covariance of local gyrification indices between bilateral paracingulate sulcus areas was calculated through partial correlations and compared by one-way analysis of variance (ANOVA) with 5000 permutations and posthoc tests. Although the reduction in covariance of patients with auditory hallucinations (PSD with H) was borderline (P = .05), subgroup analysis revealed significantly decreased covariance in patients with bipolar spectrum disorders (BSD with H) (F (2,625) = 3.19, P = .013), consistent with findings from asymmetry analyses. Note: Asterisks (*) denote statistical significance levels of the posthoc Turkey HSD test after one-way ANCOVA or ANOVA with 5000 permutations (*P < .05, **P ≤ .01, ***P ≤ .001). Psychotic spectrum disorders (PSD) encompass schizophrenia spectrum disorders (SSD) and bipolar spectrum disorders (BSD). Patients have either experienced auditory hallucination during their lifetime (H) or have not (NH). Healthy controls are denoted as (Control). Covariates include sex, age, estimated total intracranial volume, the difference between bilateral brain volumes (Left brain volume − Right brain volume), the sum of bilateral metrics (Left + Right), and broad diagnostic groups (SSD, BSD, or Control) for PSD comparisons.

The asymmetry in PCS length, as measured by the classical asymmetry index, showed consistent group differences, with F(2,626) = 5.06 and P = .002 (see Figure S2). When additional covariates, such as antipsychotic dose or PANSS positive scores, were included in the analysis, they did not change the observed reduction in PCS length asymmetry among patients with lifetime AH compared to those without (post-hoc test: M = −11.1 and −10.2 mms, Z = −2.54 and −2.36, SE = 4.40 and 4.34, *P* = .011 and .018).

#### Unilateral Analysis

In the unilateral analyses, no significant group differences were observed in either the unilateral rating scale or PCS length or depth metrics (see Figure S2 and Table S1).

### Interhemispheric covariance of PCS in patients with auditory hallucinations

To further examine sulcal pattern coordination, we calculated the interhemispheric covariance of LGIs between bilateral PCS areas, compared with healthy controls and patients with lifetime AH, patients without such history demonstrated the strongest covariances (F (2,625) = 8.38, *P* < .001). In the subgroup analysis, an increased covariance in both patient subgroups compared to controls were found, only patients with the bipolar spectrum and lifetime AH displayed significantly decreased covariance (F (2,625) = 19.7, *P* < .001; post-hoc test, M = −0.18, Z = −6.16, SE = 0.03, *P* < .001). Summarized results from interhemispheric covariance in PCS are depicted in Figure 3B and Table S2.

### Changes in interhemispheric covariance within and between salience and auditory networks

As in the PCS regions, patients exhibited significantly increased covariance between bilateral salience networks consistent in the overall group and subgroups (F (2,625) = 102 to 299, *P* < .001). In addition, the presence of lifetime AH influenced covariance differently across patient subgroups (see Figure 4A). Patients with schizophrenia spectrum disorders and lifetime AH showed increased covariance compared to those without hallucinations (post-hoc test, M = 0.12 mm, Z = 17.3, SE = 0.01, *P* < .001), while patients within the bipolar spectrum disorders and lifetime AH displayed decreased covariance compared to patients without hallucinations (post-hoc test, M = −0.03, Z = −4.82, SE = 0.01, *P* < .001). Within bilateral auditory networks (see Figure 4B), patients with lifetime AH exhibited increased covariance compared to those without lifetime AH consistent in the overall group and subgroups (F (2,625) = 23.5 to 42.5, *P* < .001). Post-hoc tests indicated mean differences ranging from 0.03 to 0.06 (Z = 1.23 to 13.5, SE = 0.01, all *P* < .003). Similar trends were observed between the bilateral salience-auditory networks as those seen in the bilateral salience networks shown above (see Figure 4C). Detailed results from interhemispheric covariance within and between salience and auditory networks are provided in Table S2.

**Figure 4.**
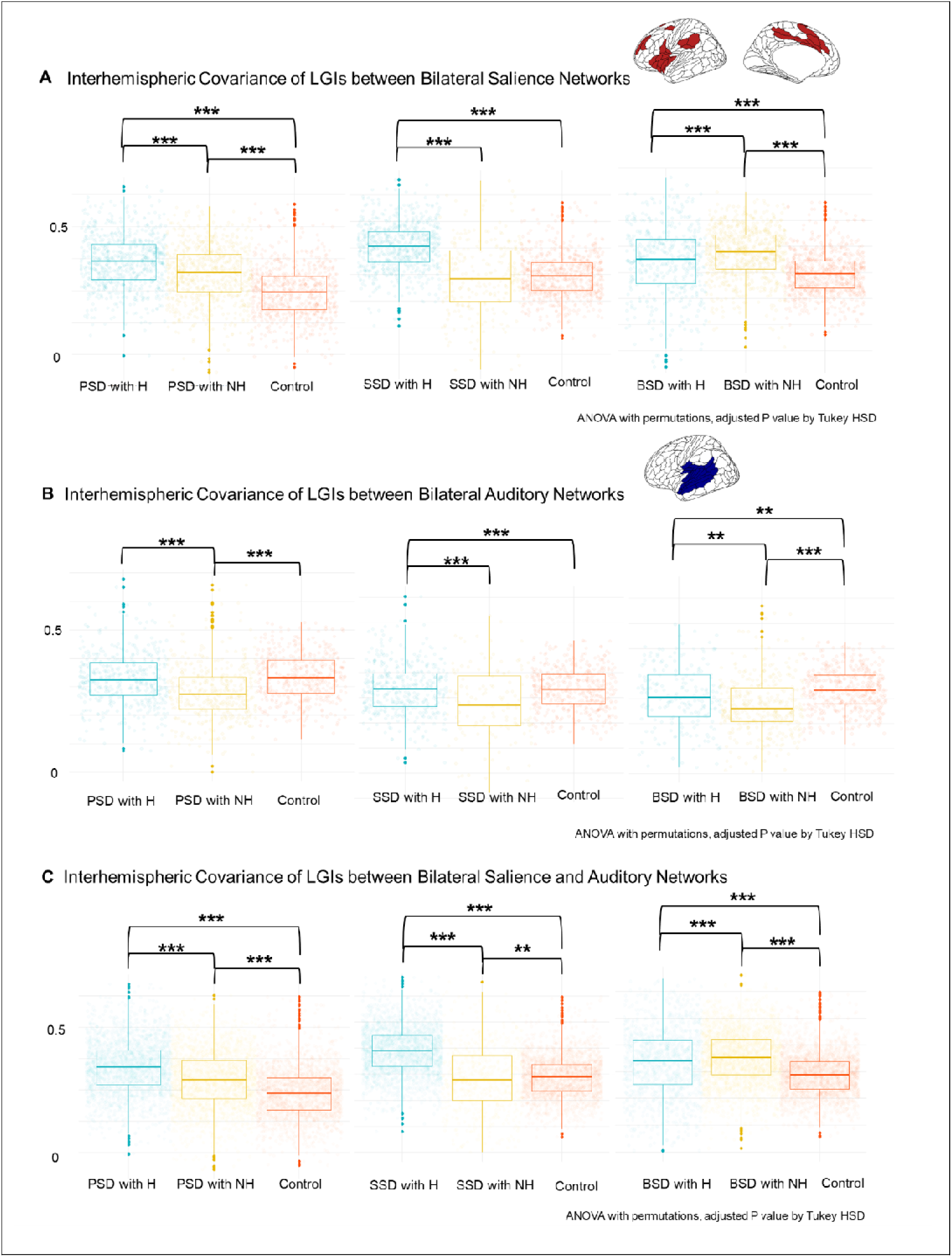
Disrupted Interhemispheric Structural Covariance of Local Gyrification Indices (LGIs) within and between Salience and Auditory Networks. (A) Patients with lifetime auditory hallucinations (PSD with H) exhibited increased interhemispheric covariance in the salience network compared to those without hallucinations (PSD with NH), while patients with bipolar spectrum disorders (BSD with H) showed decreased covariance (M = −0.03, SE = 0.01, P < .001). (B) Patients with auditory hallucinations (PSD with H) consistently showed increased covariance in the auditory network (F (2,625) = 23.5 to 42.5, P < .001). (C) Similar to the salience network, patients with auditory hallucinations (PSD with H) demonstrated increased covariance between the salience and auditory networks, while patients with bipolar spectrum disorders (BSD with H) exhibited decreased covariance (M = −0.02, SE = 0.003, P < .001). Note: Asterisks (*) indicate statistical significance levels of posthoc Tukey HSD tests after one-way ANOVA with 5000 permutations (*P < .05, **P ≤ .01, ***P ≤ .001). Psychotic spectrum disorders (PSD) encompass schizophrenia spectrum disorders (SSD) and bipolar spectrum disorders (BSD). Patients have either experienced auditory hallucination during their lifetime (H) or have not (NH). Healthy controls are denoted as (Control). Covariates include sex, age, estimated total intracranial volume, and broad diagnostic groups (SSD, BSD, or Control) for PSD comparisons.

## Discussion

The study examined PCS topological coordination in patients with psychosis, using the largest sample size to date and expanding the scope of previous research by including affective and nonaffective psychosis in the analysis and using the interhemispheric structural covariance without unilateral within-network covariance. Aligning with our hypothesis and previous studies, we observed a loss of PCS class asymmetry, replicated the decreased leftward PCS length asymmetry, and identified increased structural covariance of gyrification within and between the salience and auditory networks in patients with schizophrenia spectrum disorders and lifetime AH than those without ^5,11,50,51^. Similarly, this reduced leftward PCS asymmetry extended to the patients with bipolar spectrum disorders and lifetime AH. However, there were no group differences in unilateral PCS class or metrics, and no interaction effect of lifetime AH and sex on PCS asymmetry. Despite consistent increased interhemispheric covariance in the auditory network among patients with lifetime AH than those without, we observed opposing changes within the salience network between patients with and without lifetime AH, suggesting nuanced differences in sulcal pattern coordination across the spectrum diagnoses.

### Consistent reduction in leftward asymmetry of PCS length across the psychosis spectrum with non-replicable unilateral findings

We observed a decrease in leftward asymmetry in the length of the PCS in patients with psychotic spectrum disorders and lifetime AH. This reduction was consistent across schizophrenia and bipolar spectrum disorders, with a pronounced effect in bipolar spectrum disorders, where a shift to rightward asymmetry was observed. Similar results were obtained using other two methods. The categorical rating scale detected the absence of asymmetry in patients with lifetime AH compared to those without lifetime AH and healthy controls, with a significant impact in bipolar spectrum disorders. However, this categorical method did not detect differences between groups, possibly due to its lower sensitivity compared to continuous approaches or potential bias from directly subtracting ordinal variables. Additionally, we identified a significant decrease in the interhemispheric covariance of gyrification between bilateral PCS areas in patients with bipolar spectrum disorders and lifetime AH, compared with those without lifetime AH and healthy controls, not in patients with schizophrenia spectrum disorders.

However, we did not replicate any between-group differences in unilateral PCS measures^8–11,22–26^, whether using the categorical (Figure S1) or continuous approaches (Table S1). Several factors may contribute to this lack of replication. First, there are inconsistencies in the definition of hallucination across studies, including distinction between trait versus status, auditory versus general hallucinations, and hallucinatory perception versus behaviors. In this study, as in some studies^8–10^, we used the presence of lifetime AH from the SCID-IV, focusing on the auditory hallucinatory perception trait. In contrast, other studies often used the item P3 of the PANSS score, which reflects the general hallucinatory behaviors status^11^, or the auditory hallucinations severity^22^. Second, even when studies use similar definitions, studies focusing on the cingulate folding often have modest sample sizes or use different measurement tools^52^. Third, research cohorts might not be comparable, for example, our study and others focused on chronic or mixed-status patients^9,10,22,23,26^, while some studies emphasized first-episode psychosis^11,24,25^. How anatomical variations in the medial frontal lobe contribute to the liability for hallucinations remains unclear, though it is plausible that these variations influence brain network function, which may mediate this effect.

### Support the deceased asymmetry hypothesis without significant effect of sex

While our findings partially support the reduced asymmetry hypothesis in psychosis^2,3^, there are notable differences from the original hypothesis. First, unlike Dr. Crow comparing patients with schizophrenia or bipolar disorder to healthy controls, we compared patients with and without psychotic symptoms. Second, we found no significant interaction between sex and lifetime AH on PCS asymmetry metrics, which contrasts with the original hypothesis that posits a significant role for sex differences. While male patients with schizophrenia or bipolar disorder (particularly bipolar I) may have an earlier onset age than female patients^53,54^, there is no strong evidence of a sex difference in the onset age of auditory hallucinations in schizophrenia^55^, and data for bipolar disorders are limited^56^. Thus, we propose a revised hypothesis: reduced asymmetry across psychotic spectrum disorders, at least in the medial frontal lobe, is specifically related to auditory hallucinations, irrespective of sex differences. However, this revised hypothesis requires further rigorous testing and refinement in future research.

### Consistent interhemispheric structural covariance in auditory network, yet divergent patterns in salience network

Compared with patients without lifetime AH, patients with schizophrenia and bipolar spectrum disorders exhibited similar interhemispheric structural covariance patterns in auditory networks. However, contrasting patterns in salience networks and between auditory and salience networks were observed. This finding in the auditory network aligns with previous studies from the same cohort, which identified deviations of cortical thickness in the Heschl’s gyrus within the auditory network shared by patients with schizophrenia and bipolar spectrum and lifetime AH compared to those without such lifetime AH^37,38^. Additionally, a similar pattern was seen in high-risk patients transitioning to psychosis^30^. This increased covariance may relate to heightened temporal hyperactivation, which is critical for auditory hallucinations^57^. However, when examining structural covariance in the salience networks and between salience and auditory networks, we found divergent effects between two disorders. Patients with schizophrenia spectrum disorders and lifetime AH demonstrated increased covariance than those without lifetime AH, while patients with bipolar spectrum disorders and lifetime AH exhibited decreased covariance than those without lifetime AH, possibly linked to the hypoactivity in the salience network^58,59^. Further research is needed to elucidate the relationship between structural covariance in cortical patterns and functional connectivity.

### Recommendations

Moving forward, several recommendations for future asymmetry studies emerge. First, while our study focused on PCS asymmetry for the auditory hallucination trait, exploring other morphometric markers for psychosis status, such as asymmetry in surface area, could offer valuable insights. Second, conducting a comprehensive analysis of whole-brain structural asymmetry, encompassing subcortical structures, through both region-based and vertex-wise approaches, is pivotal for a more profound comprehension of the psychosis continuum. Third, considering the importance of cytoskeleton-related genes in brain lateralization among nonclinical populations^60^, investigating whether these genes exert influence across the psychosis continuum is imperative. Last, examining brain lateralization in high-risk subclinical adolescents with distressing psychotic-like experiences^61^ can yield valuable insights into the neurodevelopmental trajectory of the full psychosis continuum model.

## Conclusions and implications

This study, featuring the largest cohort to date investigating cingulate folding, supports the hypothesis of reduced asymmetry within the psychosis continuum. We observed consistent reduction in leftward asymmetry in the PCS among patients with psychotic spectrum disorders and lifetime AH, suggesting that the PCS morphology is transdiagnostically linked to the auditory hallucination trait. Rather than applying this theory to specific disorders, we propose that focusing on symptoms may be key to generalizing this hypothesis. Additionally, our findings highlight the importance of interhemispheric cortical coordination in the psychosis continuum, while acknowledging the complexities in specific sulcal patterns across different psychopathologies.

## Data Availability

All data produced in the present study are available to Lynn Morch-Johnsen and Ingrid Agartz upon reasonable request.

## Acknowledgments

We would like to express our gratitude to the Thematically Organized Psychosis Study group and all participants in this study, without whom this work would not have been possible. We would also like to thank Professor Paul Fletcher and Professor Peter Jones for their advice.

## Disclosures

IA has received speaker’s honoraria from Lundbeck. GKM consults for ieso Digital Health. All other authors declare no conflicts of interest to disclose.

Some contents of this manuscript were presented at the 2024 Congress of the Schizophrenia International Research Society.

