## Supplemental Tables and Figures for "Reassessing Asymmetry Reduction in Psychosis: Cingulate Folding and Gyrification Covariance in Patients with Auditory Hallucinations"

**Supplementary Materials**

**Supplementary Results**

Table S1. Paracingulate Sulcus Metrics and Asymmetry Differences

Table S2. Interhemispheric Structural Covariances of Local Gyrification Index

Figure S1. Decreased Leftward Asymmetry Index of the Length of Paracingulate Sulcus in Patients with Lifetime Auditory Hallucinations

Figure S2. Unilateral Paracingulate Sulcus Class

**Table S1. Paracingulate Sulcus Metrics and Asymmetry Differences (mean and standard deviation in mm)**

| **Sulcal metrics** **and cortical thickness of the paracingulate sulcus** | | | | | | | | |
| --- | --- | --- | --- | --- | --- | --- | --- | --- |
|  | **PSD with H** | **PSD with NH** | **Control** | **F (2,626)** | **p-val** | **p1^adj^** | **p2^adj^** | **p3^adj^** |
| **Left length** | 37.8(27.4) | 41.0 (29.5) | 41.5 (29.5) | 1.06 | 0.14 | n.s. | n.s. | n.s. |
| **Right length** | 34.0 (27.2) | 30.5 (28.4) | 32.0 (27.7) | 1.70 | 0.07 | n.s. | n.s. | n.s. |
| **Length AD** | 3.7 (34.7) | 10.5 (36.6) | 9.5 (37.1) | 3.19 | **0.01** | n.s. | 0.05 | **0.03** |
| **Left depth** | 5.7 (2.9) | 5.7 (2.6) | 6.0 (2.9) | 0.01 | 0.87 | n.s. | n.s. | n.s. |
| **Right depth** | 5.3 (2.6) | 4.8 (3.0) | 4.9 (2.8) | 0.20 | 0.08 | n.s. | n.s. | n.s. |
| **Depth AD** | 0.4 (3.9) | 0.8 (3.4) | 1.1 (3.7) | 1.03 | 0.15 | n.s. | n.s. | n.s. |
|  | **SSD with H** | **SSD with NH** | **Control** | **F (2,626)** | **p-val** | **p1^adj^** | **p2^adj^** | **p3^adj^** |
| **Left length** | 39.5 (27.9) | 44.7 (30.8) | 41.5 (29.5) | 0.37 | 0.69 | n.s. | n.s. | n.s. |
| **Right length** | 33.2 (28.2) | 28.3 (23.2) | 32.0 (27.7) | 0.87 | 0.43 | n.s. | n.s. | n.s. |
| **Length AD** | 6.3 (35.5) | 16.3 (33.4) | 9.5 (37.1) | 1.41 | 0.25 | n.s. | n.s. | n.s. |
| **Left depth** | 5.7 (2.9) | 6.0 (2.3) | 6.0 (2.9) | 0.19 | 0.84 | n.s. | n.s. | n.s. |
| **Right depth** | 5.2 (2.7) | 4.9 (2.6) | 4.9 (2.8) | 1.13 | 0.32 | n.s. | n.s. | n.s. |
| **Depth AD** | 0.5 (3.9) | 1.1 (3.2) | 1.1 (3.7) | 1.21 | 0.30 | n.s. | n.s. | n.s. |
|  | **BSD with H** | **BSD with NH** | **Control** | **F (2,626)** | **p-val** | **p1^adj^** | **p2^adj^** | **p3^adj^** |
| **Left length** | 32.4 (25.1) | 39.3 (28.9) | 41.5 (29.5) | 0.97 | 0.39 | n.s. | n.s. | n.s. |
| **Right length** | 36.4 (23.9) | 31.5 (30.6) | 32.0 (27.7) | 1.49 | 0.23 | n.s. | n.s. | n.s. |
| **Length AD** | -4.0 (31.1) | 7.8 (37.8) | 9.5 (37.1) | 2.82 | 0.06 | **0.04** | n.s. | n.s. |
| **Left depth** | 5.6 (3.0) | 5.6 (2.7) | 6.0 (2.9) | 0.35 | 0.70 | n.s. | n.s. | n.s. |
| **Right depth** | 5.6 (2.1) | 4.9 (3.2) | 4.9 (2.8) | 2.49 | 0.09 | n.s. | n.s. | n.s. |
| **Depth AD** | 0.0 (3.7) | 0.7 (3.5) | 1.1 (3.7) | 2.17 | 0.12 | n.s. | n.s. | n.s. |

SSD = schizophrenia spectrum disorders, BSD = bipolar spectrum disorders, PSD= psychotic spectrum disorders, H = lifetime auditory hallucinations, NH = no lifetime auditory hallucinations, SD = standard deviation, AD = asymmetry difference (Left - Right), thickness = cortical thickness around the PCS, F = one-way ANCOVA with 5000 permutations, p1^adj^ = H vs. control; p2^adj^ = NH vs. control; p3^adj^ = H vs. NH, n.s. = non-significance. Covariates in ANCOVA include sex, age, estimated total intracranial volume, the difference between bilateral brain volumes (Left brain volume – Right brain volume), the sum of bilateral metrics (Left + Right), and broad diagnostic groups (SSD, BSD, or Control) for PSD comparisons.

**Table S2. Interhemispheric Structural Covariances of Local Gyrification Index (mean and standard deviation)**

| **Interhemispheric structural covariances of local gyrification index** | | | | | | | | |
| --- | --- | --- | --- | --- | --- | --- | --- | --- |
|  | **PSD with H** | **PSD with NH** | **Control** | **F (2,625)** | **p-val** | **p1^adj^** | **p2^adj^** | **p3^adj^** |
| **PCS** | 0.35 (0.11) | 0.38 (0.09) | 0.32 (0.09) | 8.38 | <0.001 | 0.05 | <0.001 | n.s. |
| **Salience** | 0.39 (0.08) | 0.35 (0.09) | 0.30 (0.08) | 233 | <0.001 | <0.001 | <0.001 | <0.001 |
| **Auditory** | 0.33 (0.09) | 0.28 (0.09) | 0.33 (0.08) | 42.5 | <0.001 | n.s. | <0.001 | <0.001 |
| **Salience-Auditory** | 0.37 (0.08) | 0.33 (0.09) | 0.29 (0.08) | 587 | <0.001 | <0.001 | <0.001 | <0.001 |
|  | **SSD with H** | **SSD with NH** | **Control** | **F (2,625)** | **p-val** | **p1^adj^** | **p2^adj^** | **p3^adj^** |
| **PCS** | 0.39 (0.11) | 0.42 (0.14) | 0.32 (0.09) | 16.4 | <0.001 | <0.001 | <0.001 | n.s. |
| **Salience** | 0.41 (0.09) | 0.29 (0.13) | 0.30 (0.08) | 299 | <0.001 | <0.001 | n.s. | <0.001 |
| **Auditory** | 0.33 (0.10) | 0.27 (0.14) | 0.33 (0.08) | 23.5 | <0.001 | n.s. | <0.001 | <0.001 |
| **Salience-Auditory** | 0.39 (0.09) | 0.28 (0.13) | 0.29 (0.08) | 849 | <0.001 | <0.001 | 0.002 | <0.001 |
|  | **BSD with H** | **BSD with NH** | **Control** | **F (2,625)** | **p-val** | **p1^adj^** | **p2^adj^** | **p3^adj^** |
| **PCS** | 0.19 (0.18) | 0.37 (0.12) | 0.32 (0.09) | 19.7 | <0.001 | <0.001 | 0.01 | <0.001 |
| **Salience** | 0.34 (0.14) | 0.37 (0.10) | 0.30 (0.08) | 102 | <0.001 | <0.001 | <0.001 | <0.001 |
| **Auditory** | 0.30 (0.13) | 0.27 (0.11) | 0.33 (0.08) | 33.6 | <0.001 | 0.002 | <0.001 | 0.003 |
| **Salience-Auditory** | 0.33 (0.13) | 0.35 (0.10) | 0.29 (0.08) | 244 | <0.001 | <0.001 | <0.001 | <0.001 |

SSD = schizophrenia spectrum disorders, BSD = bipolar spectrum disorders, PSD = psychotic spectrum disorders, H = lifetime auditory hallucinations, NH = no lifetime auditory hallucinations, SD = standard deviation, PCS = paracingulate sulcus, Salience= salience network, Auditory = auditory network, F = one-way ANOVA with 5000 permutations, p1^adj^ = H vs. control; p2^adj^ = NH vs. control; p3^adj^ = H vs. NH; Covariates in partial correlations include sex, age, estimated total intracranial volume, and broad diagnostic groups (SSD, BSD, or control) for PSD comparisons.

**Figure S1.**

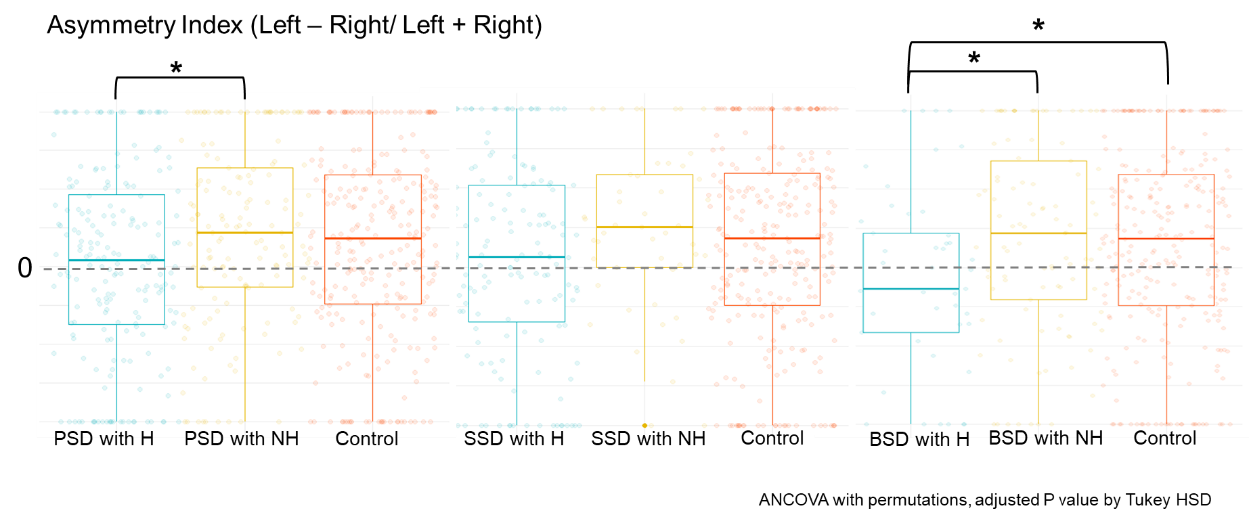

**Figure S1. More Prominent and Consistently Decreased Leftward Asymmetry Index of the Length of Paracingulate Sulcus in Patients with Lifetime Auditory Hallucinations.** We conducted an additional analysis on the asymmetry of the length of the paracingulate sulcus using the asymmetry index formula: (Left - Right) / (Left + Right), aiming to compare our findings with previous studies. This analysis yielded results consistent with our primary analysis, showing a significant decrease in leftward asymmetry, with F(2,626) = 5.06 and *P* = .002

**Figure S2.**

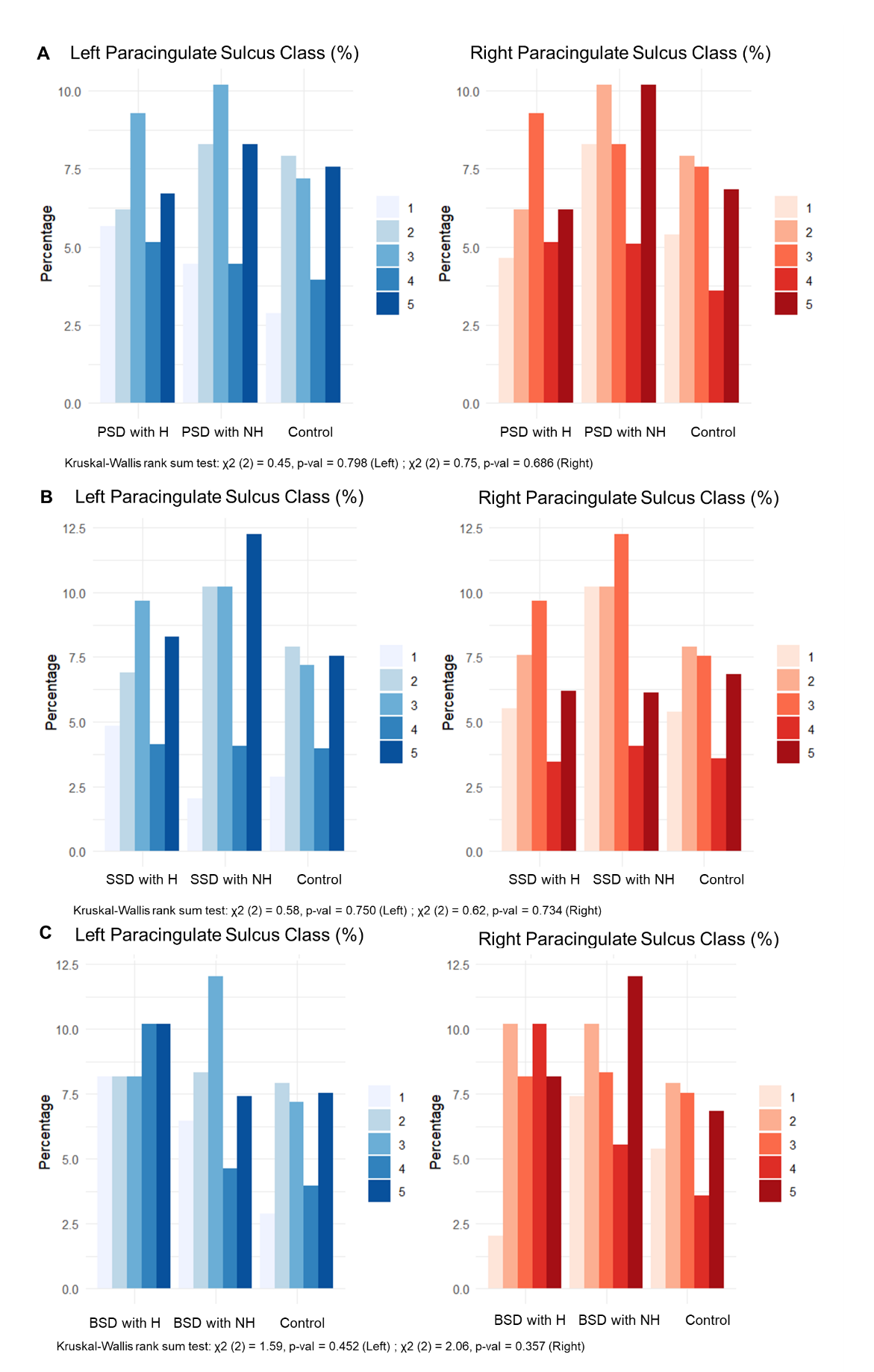

**Figure S2. Unilateral Paracingulate Sulcus (PCS) Class.** There were no significant differences between groups in either left PCS class (left side) or right PCS class (right side) among patients with auditory hallucinations (H) and those without hallucinations (NH) in (A) psychotic spectrum disorder (PSD), (B) schizophrenia spectrum disorder (SSD), (C) bipolar spectrum disorder, and healthy controls (Ctrl).
